# Polarizer-assisted pupillometry through closed eyelids overcoming pupil position dependence

**DOI:** 10.1101/2025.07.27.25331969

**Authors:** Michal Tepper, Omer Ben Barak-Dror, David Haggiag, Israel Gannot, Yuval Nir

## Abstract

Accurate assessment of pupil dynamics and gaze direction is vital in both clinical and research settings. However, conventional pupillometry methods typically require open eye conditions, which limit their applicability in patients under sedation, anesthesia, or during sleep. We previously introduced a novel, touchless short-wave infrared (SWIR) imaging method capable of separately monitoring pupil size or gaze direction through closed eyelids. Yet, clinical bedside applications demand a more robust approach that performs pupillometry while gaze direction changes dynamically, when both eyes are closed, and yields improved signal-to-noise ratio (SNR). Here, we expand upon the initial proof-of-concept by incorporating polarizing filters into the SWIR imaging setup to enhance contrast and signal quality. Multiple polarizer configurations, including crossed and parallel setups, were evaluated in both open- and closed-eye conditions to assess their effects on SNR during pupillary light reflex (PLR) imaging. In addition, we implemented improved image-processing algorithms to enable accurate pupil localization and gaze direction estimation under naturally closed eyelid conditions. We find that our improved setup employing crossed polarizer configuration significantly improved PLR SNR compared to a setup without polarizers, due to improved pupil localization and reduced surface glare. These findings suggest that the integration of polarizers into SWIR-based pupillometry systems enables more accurate and reliable measurements in clinically relevant scenarios where eyes remain closed. This advancement opens opportunities for broader applications in sleep medicine, anesthesiology, and neurocritical care.

## Introduction

Pupillometry, the measurement of pupil size and reactivity, is a valuable tool in various fields, including neuroscience, cognitive research, and clinical medicine. The muscles controlling pupil diameter are regulated by the autonomic nervous system, causing pupil constriction and dilatation. Pupil dynamics occur in response to changes in ambient light, fixation, following transient external stimuli such as light or sound, activation of neuromodulatory pathways such as noradrenaline signaling^1–3^, or due to internal factors including arousal, pain, cognition, and mental effort^4–6^.

Modern pupillometry methods are used for a variety of clinical applications and have evolved from manual measurement using a penlight and a ruler to automated devices measuring and tracking pupil size and position over time^7^. In medicine, pupillometry is used as a non-invasive tool to detect abnormalities in autonomic function, which can indicate issues such as traumatic brain injury^8^, stroke^9^, intracranial pressure^10^, neurodegenerative diseases^11^, diabetes complications^12^, and brainstem function impairment^7,13,14^. Given its tight link with arousal, pupil size also can be a useful marker for anesthesia depth^15,16^ and sleep stages^17,18^. In psychology, pupillometry is used to study emotional and cognitive responses, enhancing the study of human behavior and mental processes^5,19,20^. The correlation of pupil dynamics to internal cognitive and physical processes makes it a valuable tool for biofeedback applications^21,22^, as well as an additional input source for brain-computer interface (BCI) systems^23,24^.

Despite the ongoing research and development of quantitative automated techniques and methods, clinical pupillometry requires a human operator, making the measurements time-consuming, intermittent, and less accurate^25^. Manual measurements, as well as automated devices, rely on viewing the pupil in visible or near infrared wavelengths. Since the eyelid is relatively opaque in these wavelengths, these pupillometry devices can only be used in open-eye situations. Some attempts have been made in closed eye pupillometry in specific species such as pigeons where the eyelid is translucent^26^, or in humans when delivering light through the temple to “back-illuminate” the eye, but this technique has limited applicability as it requires frequent breaks to avoid heat damage to the tissue^27^. Thus, despite its clinical potential, pupillometry in its current form cannot be employed to continuously monitor closed-eye situations such as surgical anesthesia or sleep.

We recently presented a method enabling continuous touchless pupillometry in closed eyes using short-wave infrared (SWIR) imaging and dedicated algorithms^28^. Since SWIR wavelengths can penetrate through the eyelid skin, SWIR imaging can provide visualization of the iris and pupil behind it^29^. The original proof-of-concept monitored gaze direction and pupil size changes induced by the pupillary light reflex (PLR) in highly controlled settings when participants directed their gaze forward and held one eye closed. To move towards clinical bedside applications, we opted to track pupil size while eye gaze and pupil position change freely, when both eyes are naturally closed, and we aimed to increase its performance and improve its signal-to-noise (SNR). One of the main contributors to the noise in the SWIR image of the closed eye is the reflected light, or glare, from the eyelid skin surface. Eliminating this glare is desirable to improve visualization of light scattered from the pupil and iris.

One of the most widely used optical techniques to improve tissue visualization is polarization imaging. It uses polarizers which are optical elements that selectively transmit light waves with a specific polarization orientation while blocking others. By selectively filtering polarized light, polarizers aid in visualizing structures with greater clarity and detail. When integrated into optical devices, polarizers can improve diagnostic accuracy and facilitate insights into deeper biological processes^30^. In biomedical imaging, polarizers play a critical role in enhancing contrast, reducing glare, and improving image quality in modalities such as microscopy^31^, optical coherence tomography (OCT)^32^, biomedical polarization imaging^33^, and fluorescence imaging^34^. Since the polarization of reflected or emitted light depends on the structure of the observed object, the use of polarizers enables researchers to selectively enhance or minimize the visibility of certain features.

In addition, the use of a crossed-orientation polarizer configuration (one polarizer between the illumination source and the tissue, and another polarizer with orthogonal orientation placed between the tissue and the camera) can assist in removing surface glare from the illuminated tissue. When the tissue is illuminated, the light is partially reflected from the tissue surface, while relatively maintaining the original polarization. Most of the incident light does not undergo surface reflection but rather penetrates the tissue, where it is subjected to multiple scattering events by heterogeneous structures. These interactions result in depolarization, such that the emerging light exhibits a randomized polarization distribution. As a result, the second polarizer placed before the detector transmits approximately half of the depolarized light, thus permitting meaningful image formation even in a crossed setup, while suppressing a substantial portion of the surface glare.

When imaging the eye, polarized imaging of the iris and cornea assists clinicians to gather valuable information about tissue composition and pathological changes, aiding in the early detection and monitoring of diseases like glaucoma or malaria^35–39^. Polarimetric imaging of the eye is influenced by the birefringence properties of the iris, which alter the polarization of transmitted light. This variability complicates the determination of the optimal polarizer configuration for polarization-based eye imaging devices^40,41^. While polarizers could potentially reduce the noise and improve the accuracy of closed eye pupillometry, they also reduce signal amplitude as they decrease light intensity reaching the camera. Thus, a detailed examination is necessary to determine if and how polarization imaging could improve closed eye pupillometry and gaze direction estimation. To this end, we investigated polarized imaging in open eyes settings to inform subsequent experiments with closed eyes. The study also implemented further improvements to closed eye imaging via automatic identification of pupil position in closed eyes that enables pupillometry in the face of ongoing eye movements, and by transitioning to naturally closed eyes settings.

## Methods

### Study Design

Experiments were conducted on healthy volunteers in accordance with the institutional review board of the ethical committee Tel Aviv University (approval no. 0005827-1) and radiation safety requirements (IEC 62471 international standard). Each volunteer signed an informed consent for participating in the study. The experiments were based on artificially inducing changes in pupil size (in response to brief light stimuli) and pupil position (directing gaze to specific directions in response to instructions) and measuring the PLR in closed eye settings using the methods described below.

### Experimental setup

All experiments were conducted in a dark room. The experimental setup is shown in Figure 1. Participants sat while their head rested on a dedicated chinrest (Eyelink®, SR Research, Ottawa, Canada) with their heads faced towards a computer screen at a distance of 40 cm (Dell 27 Monitor – P2719H, typical brightness of 300 lux, Dell Inc, Round Rock, TX). They were asked to sit still and keep their eyes closed during the experiments, maintaining the same gaze direction unless requested otherwise, and were allowed at least 30 seconds to adapt to the dark before visual stimulation began. The participant’s face and eyes were illuminated with an 1100 nm LED (light emitting diode) (Thorlabs M1100D1, 168mW(min)) placed 23cm from the chinrest at the approximate height of the eyes. Both eyes were imaged using a SWIR camera (WiDy SenS 640V-ST, New Imaging Technologies (NIT), Verrières le Buisson, France) equipped with 25mm lens (LM16HC, Kowa, Dusseldorf, Germany) placed 30cm from the chinrest directly in front of the participants at the approximate height of the eyes. The video was captured using the camera’s software, at a 31Hz frame rate with a 15ms integration interval. The main experimental setup included two polarizers: one in front of the LED (referenced in this manuscript as the “LED polarizer”) and the other in front of the camera (the “camera polarizer”) (W436160 and W428316 respectively, Thorlabs, Newton, NJ).

**Figure 1:**
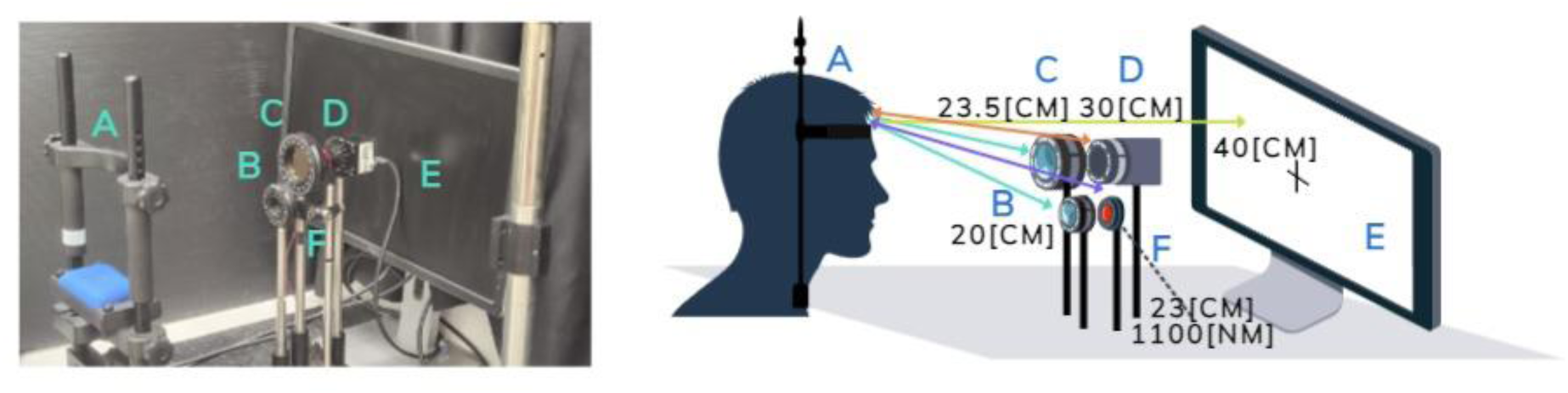
Experimental setup. A: Chinrest, B: LED polarizer, C: Camera polarizer, D: Camera, E: Screen, F: LED. The polarizers were replaced by a beam splitter at the position of C for the control configuration

To compare the results with a setting using unpolarized light, while maintaining the same illumination intensity reaching the eyes, a control experimental setup was used. In this configuration, both polarizers were removed, and a beam splitter (CM1-BS014, Thorlabs) was placed in front of the illumination LED. The beam splitter reduced the light intensity by 50%, ensuring the eyes were exposed to the same illumination intensity in all three configurations, under the assumption of polarization-independent LED illumination.

### Experimental design

#### Pupillary light reflex (PLR)

Comparing different polarizer orientations is complicated by the different characteristics of each participant and experiment. Small changes in the participant position or orientation towards the camera may lead to different illumination levels on the eyelid. Therefore, standardized stimulation was used to induce comparable responses between different polarizer configurations. In each experiment, 10 PLR responses were induced by presenting a white screen stimulus for two seconds on the computer screen in front of the participants. Visual stimulations were interleaved by periods of dark inter-stimulus-intervals lasting 20±2 seconds (to avoid prediction bias), as previously described^28^. The PLR response is a reflexive response with a characteristic time-course^42^ that is relatively consistent between experiments and could thus provide a reliable basis for comparison.

#### Experiment 1 - open eyes

We compared the PLRs in open eye settings in three different polarizer orientations: parallel orientation (both polarizers horizontally aligned), partially crossed orientation (horizontal alignment for the LED polarizer and 45° orientation for the camera polarizer), and crossed orientation (horizontal alignment for the LED polarizer and vertical orientation for the camera polarizer). The LED polarizer orientation was horizontal in all experiments as it seemed to minimize the observed reflection from the open eyelid edges.

#### Experiment 2 - forward-fixated PLR in closed eyes

The PLR experiment was conducted on six participants for each of the three configurations. The experiments were conducted using the control configuration (beam splitter without polarizers) and two polarizer configurations (in which the LED polarizer was oriented to block horizontally polarized light and the camera LED was set to either a horizontal or vertical orientation). Participants were asked to close both eyes and try to look forward (towards the estimated direction of the camera) to maintain a similar and central position of the pupil. The experiment lasted a total of ±240 seconds for each configuration.

#### Experiment 3 – PLR with varying gaze directions in closed eyes

A modified PLR experiment was conducted when the participant’s gaze was not fixated directly to the center of the camera but otherwise was arranged as the forward-fixated experiment. Here, after a 15 sec baseline period, participants were instructed to direct their gaze towards nine different directions, in set intervals, while their eyes were closed. The nine directions represented a 3*3 grid that included combinations of up-forward-bottom with left-center-right. For each direction, the participants were asked to keep their gaze fixed for 22 seconds that included 10 seconds adaptation to the new gaze direction, 2 second light stimulation using the white screen, and a 10-second darkness period. Nonuniformity in timing between PLR instances was naturally introduced due to the use of verbal instructions and the need to change gaze directions. Therefore, jitter was not incorporated into the experimental design to simplify the experimental procedure.

### Data analysis

#### Analysis of closed-eye images

PLR dynamics in closed-eye SWIR data was analyzed using the previously described ‘fixed circle’ approach^28^. In this method, the average pixel intensity is calculated within a fixed circle around the estimated pupil center with the approximate diameter of the iris. When the pupil constricts or dilates, its dark pixels occupy a different portion of the fixed circle affecting the average pixel intensity. Accordingly, we previously established that the dynamics of the intensity changes over time is similar to the changes in pupil size, facilitating the closed-eye estimation of pupil size. To ensure that changes in pixel intensity were not a result of external brightness changes, the average intensity within a control circle on a neutral area was calculated as a control. We used a white sticker placed on the participant’s forehead for the control analysis to account for brightness changes due to the participant movement, while avoiding reflection fluctuations due to involuntarily forehead muscle twitches that may occur in response to light stimulation.

For each frame of the captured video, we defined three circles: one for each eye and one as a control. Average pixel intensity data were forward-backward filtered, using a 1 Hz low pass filter (Python function “Filtfilt”^43^ - zero phase digital filter forward and backward). After this pre-processing, time courses were averaged for each participant to reflect their overall dynamics. Baseline brightness was defined as the average brightness during two seconds before light stimulation onset. Intensity dynamics were expressed in terms of pixel “darkness” changes (pixel brightness change in percentages multiplied by −1) to achieve a time-course similar to pupil size changes (the average pixel “darkness” decreases when the pupil diameter decreases shortly after light stimulation, and vice versa).

Polarizer configurations were compared using two metrics that quantify changes in brightness relative to a baseline period, defined as the two seconds preceding light stimulus onset: (1) The maximum relative change in brightness from the average baseline level, divided by the baseline brightness value, and (2) the SNR, calculated as the maximum brightness change divided by the standard deviation of the baseline brightness. These metrics were chosen for their robustness and relative insensitivity to confounding factors such as participant movement and baseline pupil size variability.

#### Analysis of open-eye images

The analysis method used for the closed eyes was used for the open eye experiment with the following modification, aimed to minimize the effect of blinks occurring intermittently in open eye conditions. Frames that included blinks were excluded and the calculated brightness was replaced by the linear extrapolation of the brightness values before and after the blinks. In cases where the blinks were at the start or end of the analysis time frame and only if the brightness reached a plateau level, a constant value with the value of the last/first non-blink frame was used to replace the blink brightness values. 4 PLR events out of 31 (12.9%) were excluded from the analysis due to excessive blinking or blinks that did not enable an accurate linear extrapolation. At least 3 PLR events per configuration for each participant were included in the analysis after this exclusion.

#### Closed-eye detection of pupil position

In closed eye experiments, especially when the participant is not looking forward, it is difficult to determine the position of the pupil. To identify the estimated position of the pupil, we used two visual aids. The first is the SWIR image of the participants eyes. As mentioned previously^28^, it is possible to see the reflection of light from the central iris and pupil as a dark circle on the eyelid (Figure 2a). Additionally, we used an image of the differences in the pixel intensity between the average intensity of two seconds baseline before the appearance of the light stimulus and one second after the stimulus onset (approximately the maximal change). Due to the short interval, the main change in the image is expected to be a result of the pupil size changes (Figure 2b). Occasionally, minimal facial movements may also be observed and can be used to assess participant movement. The diameter of the fixed circle average brightness calculation was set at 50 pixels, based on the most affected regions in these images and slightly smaller than the size of the iris.

**Figure 2:**
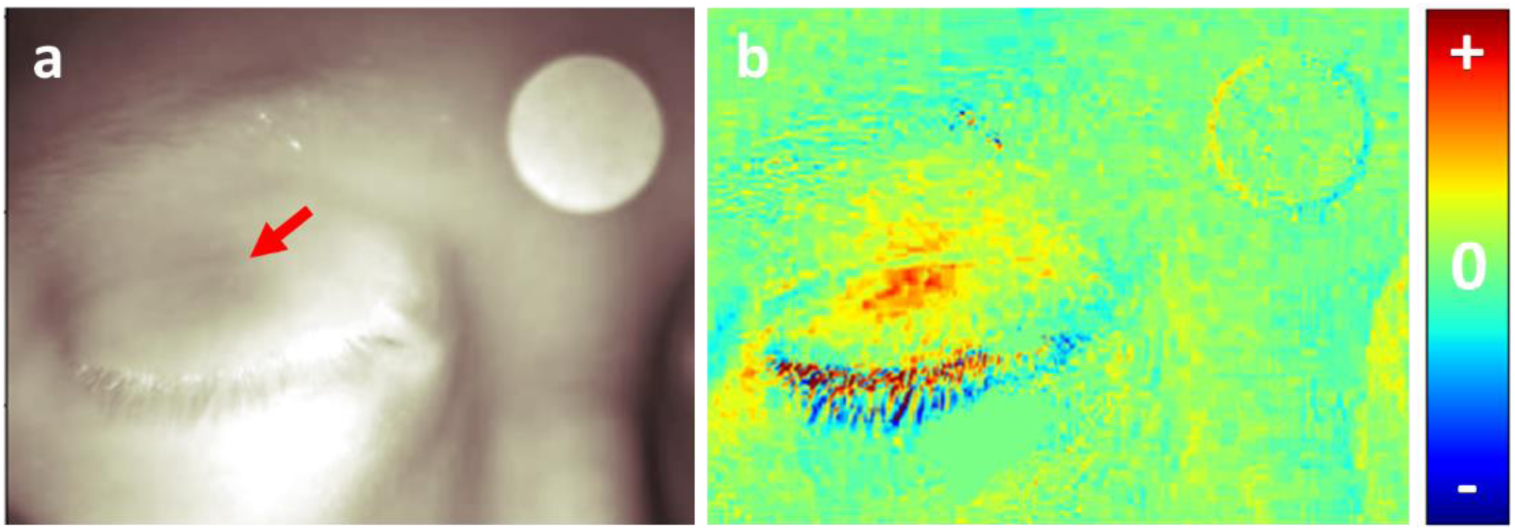
Identifying the pupil position in closed eye images. (a) original SWIR image of the participant face (captured with the crossed polarizer configuration). The light reflected from the iris can be seen as a dark region in the eyelid (marked by red arrow). (b) Brightness difference image between the average SWIR brightness image during the two seconds before the white screen illumination, and the image captured one second after the start of the white screen illumination. Red pixels in the eye reveal the area in which brightness increased corresponding to where the pupil constricted. Some movement can also be seen around the eyelashes. The brightness scale can be adjusted to exclude changes outside the eyelid area.

#### Statistical analysis

Statistical analysis was conducted in R (version 4.5.1; R Core Team 2025)^44^. A linear mixed-effects model^45,46^ was used to evaluate the effect of polarization configuration on the maximal brightness changes and the SNR. Participant identity was included as a random intercept to account for repeated measurements within individuals. Polarization configuration was modeled as a fixed effect to assess its impact on the outcome. Post-hoc pairwise comparisons between polarization configurations were performed using Tukey-adjusted tests via the multcomp package^47^ (glht()), with adjusted p-values computed using the single-step method.

## Results

To improve gaze and pupil monitoring through closed eyelids, we first investigated the utility of adding polarizers in our optical setup to eliminate glare in SWIR images. Healthy participants sat while their head rested on a dedicated chinrest and participated in experiments where brief (2sec) light stimuli were presented every 20sec on a computer screen (Methods) to induce pupillary light reflex (PLR) as described previously^28^.

### Preliminary experiments

A preliminary open-eye experiment was conducted on three participants (female in her 40s, blue eyes; female in her 30s, green eyes; male in his 20s, brown eyes) to quantify the observed changes in the brightness of open eyes during PLR response. Each experiment included at least 3 PLR events for each of the three different polarizer configurations studied. All the visual stimuli induced PLR responses for all participants. Figure 3a shows images of the eyes of all participants for the different polarizer configurations at the fully constricted pupil states. Noticeably, for all the participants, the reflection from the skin around the eyes is higher in the parallel configuration. In addition, the iris appears darker for the crossed-orientation polarization compared with the partially-crossed and parallel orientation polarizations. Since the pupil is always black in SWIR images, the brighter iris appearance led to a higher contrast between the pupil and the iris in the parallel orientation (i.e., a more visible pupil). The visual pattern within the iris seems to be reversed between the parallel and the crossed configurations. This observation (more visible pupil in parallel orientation in open eye settings) was supported by quantitative brightness changes calculated using the fixed circle approach across all PLR trials: changes were larger in amplitude in the parallel configuration throughout the PLR response (Figure 3b).

**Figure 3.**
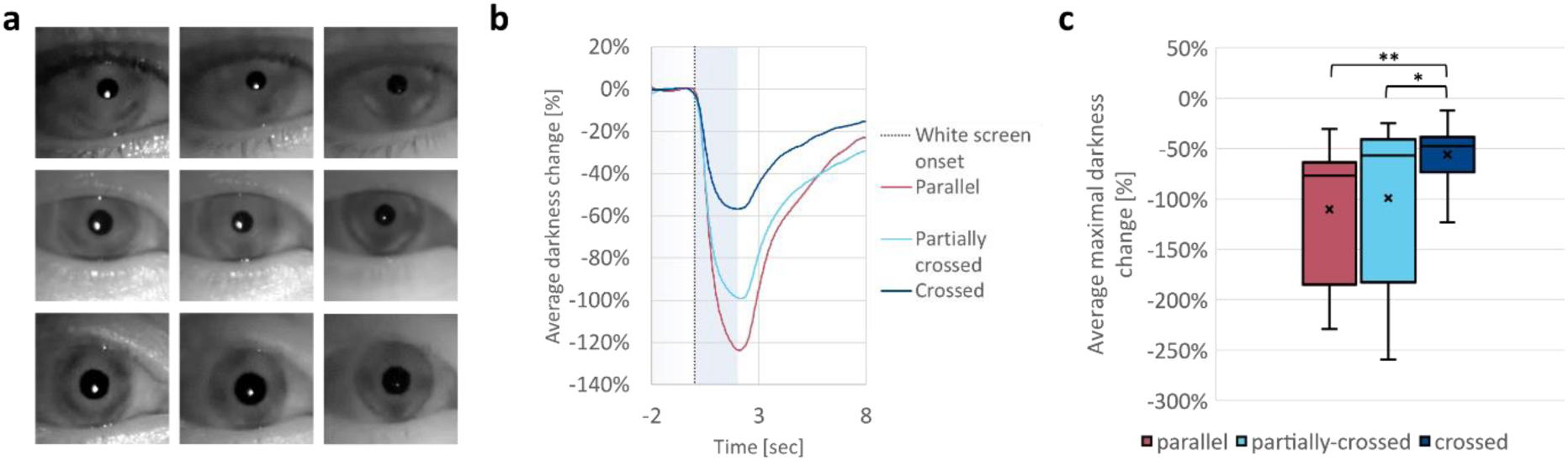
Open eye experiment. (a) Images of participant eyes at the fully constricted state for all three polarizer configurations (parallel, partially crossed and crossed orientations). The contrast between the iris and the pupil appears to be higher for the parallel configuration. (b) Grand average of all PLR events for each polarizer configuration (n= 18-24 for each polarization configuration). The 2-second duration of the white screen trigger event is marked by the light blue background. (c) Comparison of maximal average darkness changes, revealing a significantly higher change for the parallel configuration compared with the crossed configuration (p=<0.001).

We evaluated the effects of polarization configuration on both the maximal brightness change and the SNR using linear mixed-effects models with random intercepts for subjects. Maximum brightness change, expressed as a proportion of baseline, was significantly higher in the parallel configuration (M = 1.09) than in the crossed (M = 0.55; Estimate = 0.600, p < 0.001) and marginally higher than in the partially crossed configuration (M = 0.98; Estimate = 0.215, p = 0.078). The partially crossed configuration also produced significantly greater brightness than the crossed configuration (Estimate = 0.385, p = < 0.001). The distribution is shown in Figure 3c.

For SNR, the parallel configuration again yielded the highest values (M = 220), with marginally higher SNR compared to both crossed (M = 74.7; Estimate = 157.8, p = 0.093) and partially crossed (M = 65.4; Estimate = 175.9, p = 0.064) configurations. No significant difference was observed between the crossed and partially crossed configurations (p = 0.973).

These findings suggest that, in open eyes settings, while both the parallel and partially crossed configurations enhanced brightness relative to the crossed configuration, only the parallel configuration also provided an advantage in signal quality.

An opposite trend was observed when imaging closed eyes. Figure 4 shows an example of SWIR images of the same participant imaged with the three polarizer configurations. As expected, the specular reflectance from the surface of the eyelid skin was reduced with the crossed polarizer orientation. A similar reduction can also be seen in the skin and nail of the finger holding the eyelid. As these images demonstrate, there is a trade-off between better pupil contrast in the parallel configuration and the reduced noise due to the specular reflection in the crossed configuration.

**Figure 4.**
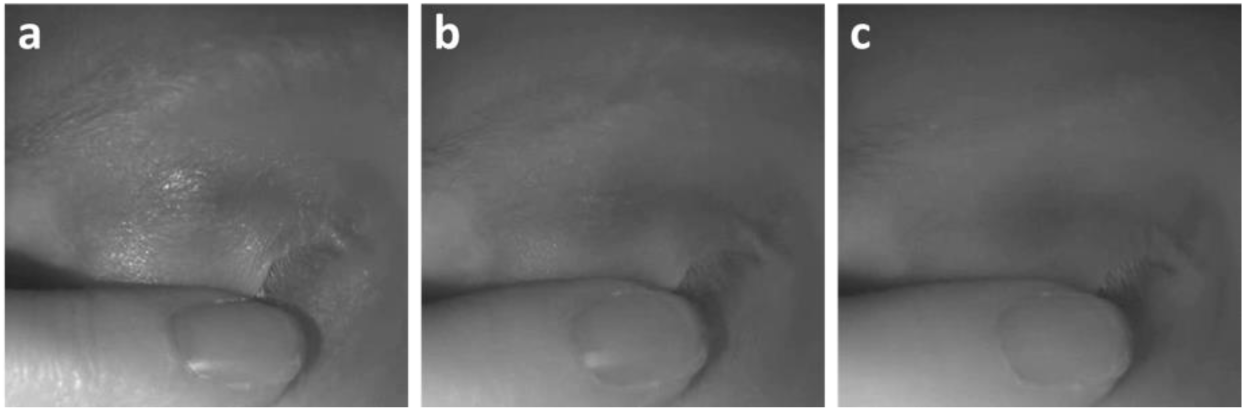
Closed eye images imaged using different polarizer configurations. (a) parallel configuration, (b) cross-polarized configuration, (c) crossed configuration.

### Forward-fixated PLR in closed eyes setting

Three polarizer configurations (parallel and crossed polarizer orientation, and the comparative no polarizer configuration, as described in the Methods section) were used to capture PLR events in 6 participants (ages: 26-41, 4 females and two males). The experiments included 10 PLR events for each participant, analyzed separately for each eye, as described above. Figure 5 displays the grand average of all PLR responses for all participants at each configuration (n=120 trials: 6 participants * 2 eyes * 10 PLR events).

**Figure 5.**
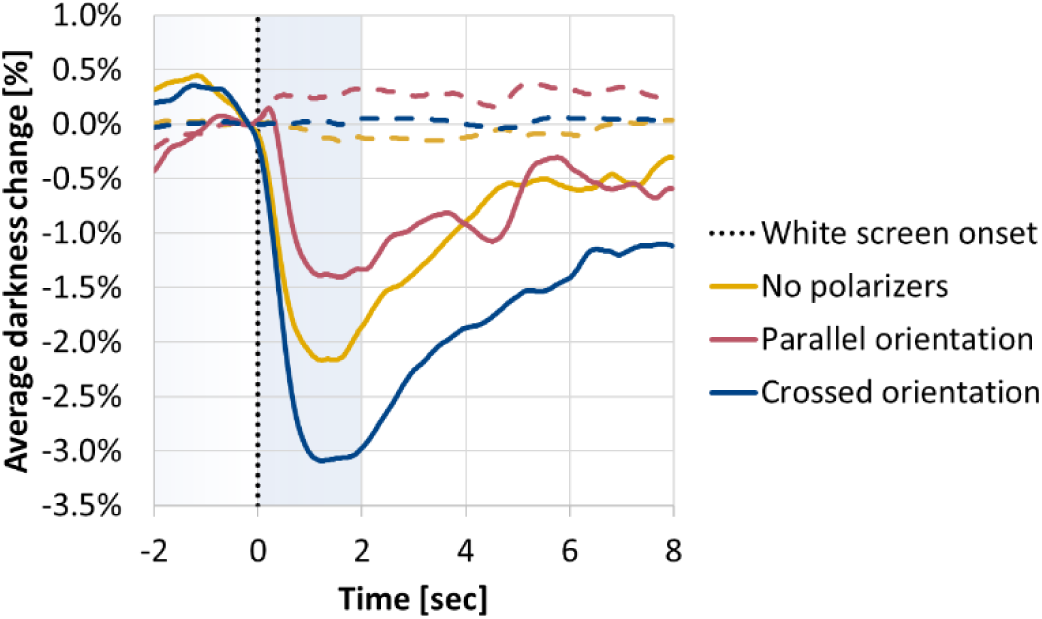
Grand average of the PLR events in the main closed-eye experiment. Continuous lines: Average darkness change for the pupil area (n=120 trials for each configuration). Dashed lines: Average darkness change for the forehead control area (n=60 trials for each configuration). Orange, no polarizers; Red, parallel polarizer orientation; Blue, crossed polarizer orientation.

The crossed orientation configuration yielded the highest mean brightness change (M = 0.0171), significantly greater than both the no-polarizer configuration (M = 0.0106; Estimate = 0.00644, p = 0.035) and the parallel configuration (M = 0.00580; Estimate = 0.01126, p < 0.0001). The difference between the parallel and no-polarizer configurations was not statistically significant (p = 0.152).

SNR followed a similar trend, with the highest value observed in the crossed configuration (M = 13.6), which was significantly greater than in the no-polarizer (M = 9.79; Estimate = 3.81, p = 0.0015) and parallel (M = 7.63; Estimate = 5.97, p < 0.001) configurations. No significant difference was found between the parallel and no-polarizer conditions (Estimate = –2.16, p = 0.118).

Together, these results show that in the closed-eye experiment, the crossed polarization configuration enhanced both brightness and signal quality compared with the no polarization or parallel orientation configurations.

### PLR with varying gaze directions in closed eyes setting

The same polarizer configurations were used to image PLR events when the participants were instructed to direct their gaze to different directions (Methods). The brightness difference and the SWIR images were used to identify the pupil for each PLR event separately as described above in Figure 2. The crossed orientation configuration enabled better identification of pupil position, since the specular reflection was very sensitive to small movements, leading to a more pronounced effects in the brightness difference images (Figure 6).

**Figure 6.**
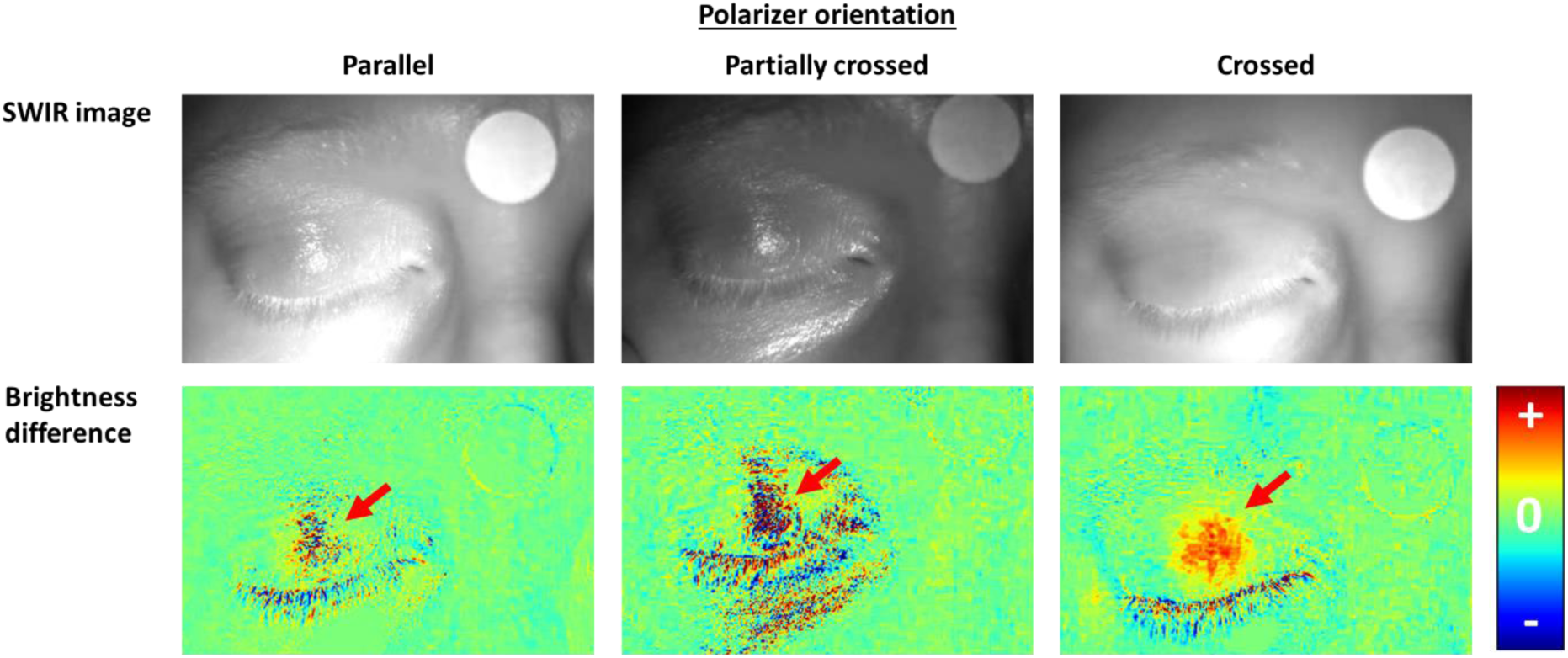
Comparing polarizer configuration to estimate pupil position in closed eye settings. Brightness difference images for the different configurations, demonstrating the added value of the polarizers in removing the glare from the eyelids and revealing the pupil position (marked by arrows) more clearly. Columns represent (left) Parallel configuration, (middle) crossed configuration, (right) control (without polarizers) configuration. Bottom row shows the brightness difference in images obtained before and after brief light stimuli were presented.

Figure 7 displays the pupil identification analysis, for all nine gaze directions, in a representative individual for the right eye only. Pupil position could be identified in each gaze direction to guide positioning the ‘fixed circle’ region-of-interest for analysis of PLR dynamics. As can be seen, the crossed orientation allowed the identification of the typical PLR responses in most gaze directions even when applied to single trials, without the need to average over multiple events.

**Figure 7.**
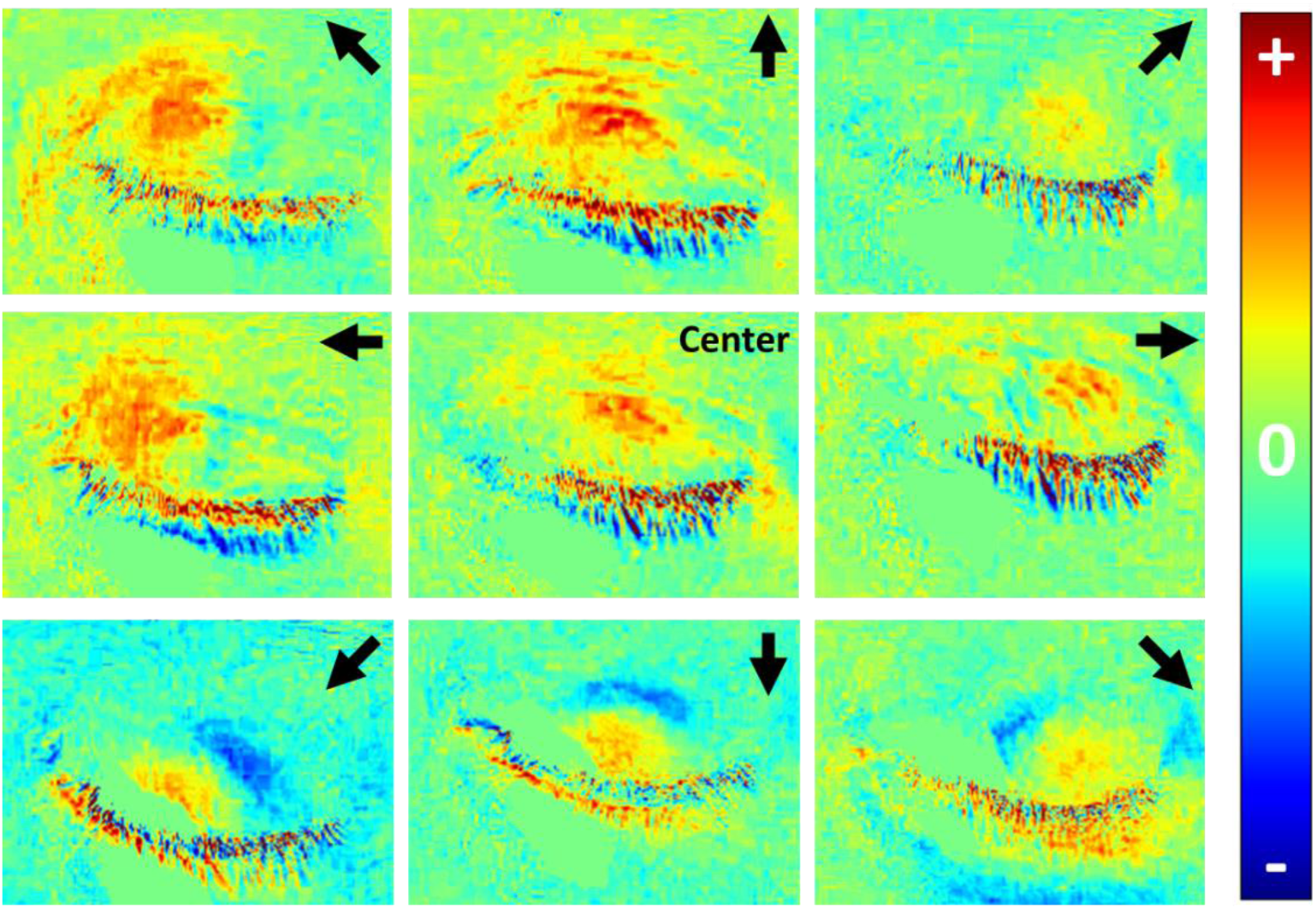
Imaging PLR events for varying gaze directions: single participant outcomes. SWIR subtraction images for nine different gaze directions. The red regions reveal areas in which the brightness increased, suggesting pupil constriction, which guided positioning of ‘fixed circle’ region of interest for subsequent analysis. The images are organized according to the instructed gaze direction (marked by black arrows or “center” at the top right corners of each image).

Next, we proceeded to quantitative analysis across the entire dataset. Figure 8 displays the outcomes of this experiment, averaged for both eyes, across all participants, and across all trials (n=108 PLR events for each polarizer configuration). The typical PLR response curves can be seen for most configurations and positions (Figure 8a) and were most prominent in the crossed orientation. Some gaze directions showed poorer results. It should be noted that some of the participants commented that it was harder to maintain a fixed stare direction in some of the gaze directions (mostly the top-left and top-right directions). In addition, the eyelashes added noise to the analysis that was more evident in the bottom gaze directions. Despite these challenges, the grand averages of the responses for each of the configurations, over all gaze directions (Figure 8b) displays a very clear PLR response for all polarizer configurations. The crossed polarizer configuration produced the highest mean brightness change (M = 0.0146), significantly greater than both the no polarizer (M = 0.00508; Estimate = 0.00957, p = 0.0079) and parallel (M = 0.00433; Estimate = 0.01031, p = 0.0036) configurations. No significant difference was observed between the parallel and no polarizer configurations (Estimate = – 0.00075, p = 0.97).

**Figure 8.**
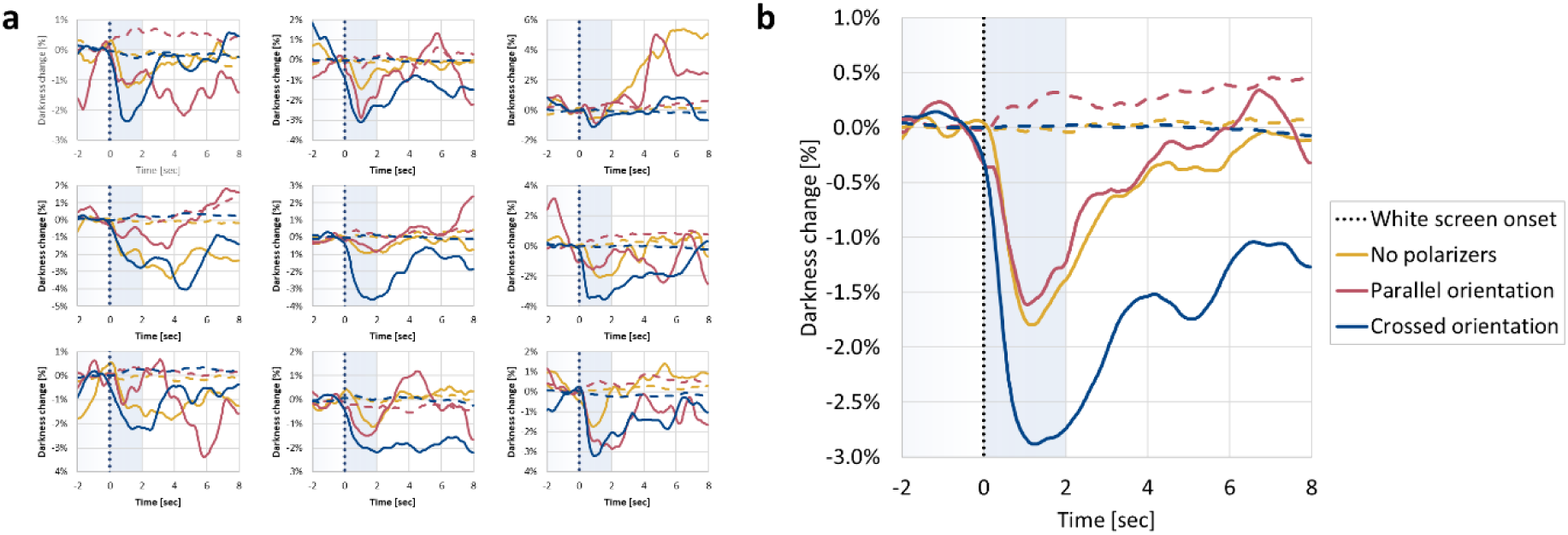
Imaging PLR events for varying gaze directions: grand average on all participants. (a) Darkness changes following the PLR events for each position, at each polarizer configuration (n=12 for each curve). (b) Grand average over all positions (n=108 for each configuration).

Similarly, SNR was highest in the crossed polarizer configuration (M = 12.1), with significantly higher values than both the no polarizer (M = 6.90; Estimate = 5.19, p < 0.001) and parallel (M = 5.78; Estimate = 6.31, p < 0.0001) configurations. Again, no significant difference emerged between the parallel and no polarizer configurations (Estimate = –1.12, p = 0.66).

The forehead region in the parallel configuration was relatively noisier than for the other configurations, further demonstrating the problematic nature of specular reflection noise in this configuration. These findings further support the conclusion that crossed polarization markedly enhances both image brightness and signal quality in the closed eye settings, while the parallel configuration does not confer improvements over the control configuration without polarizers.

## Discussion

Closed eye pupillometry has a big potential to impact multiple medical and scientific fields. In this study, we enhance the SWIR-based imaging method by introducing several key improvements over our original proof-of-concept report^28^. Specifically, we incorporate polarizers into the optical setup, develop software tools capable of tracking pupil size despite positional changes, and perform imaging with both eyes naturally closed (without manually stretching the eyelid). These refinements enable imaging under conditions that more closely resemble clinical scenarios. Despite the added challenges of imaging through thicker, naturally closed eyelids (in previous experiments, eyelid of closed eye was stretched by placing a finger on it) and the uncertainty of pupil position, the maximal pupillary light reflex (PLR) amplitude averaged approximately 2%, consistent with our previous findings.

The closed-eye experiments demonstrate a greater change in the PLR response when using the crossed polarizer configuration compared to the control setup without polarizers, mainly in the varying gaze direction experiment. This improvement is due to the reduced glare and the consequent improved pupil position estimation enabled by the cleaner image. This highlights the broader utility of polarization in biomedical imaging. By selectively filtering light based on its polarization state, the use of polarizers can enhance contrast, suppress surface reflections, and isolate subsurface scattering - features particularly beneficial for imaging through turbid or layered tissues like skin or eyelids. Polarization-based techniques are increasingly used in various biomedical applications. For example, In dermatology, polarization imaging can improve visualization of vascular structures and pigmented lesions by filtering out surface glare and enhancing contrast in melanin-rich tissues^48^. In neurosurgery, polarized-light imaging can be used to identify structures associated with cancerous tissue and assist in the detection of tumor borders^49^. Similarly, a Mueller polarimetric endoscope provided multiple polarization-contrast images and demonstrated enhanced visualization of early epithelial cancers and clear tumor margins compared to standard endoscopy^50^. Furthermore, polarization filtering can be implemented with minimal changes to existing hardware and without increasing illumination power, making it a low-cost, noninvasive strategy for enhancing signal fidelity in both research and clinical imaging systems.

The study also presented several improvements over the analysis algorithms described previously. The previous study was based on a more rigid experimental setup due to the need to establish the validity of this method and compare its pupil size and position estimations to a simultaneous open-eye control. Therefore, participants held one eye closed using their finger and kept the other eye open. As discussed in detail in the previous study, holding the eyelid does not represent clinically relevant scenarios, in which the eyes are naturally closed. In the current study all closed eye measurements were conducted with naturally closed eyes. This change necessitated accurate identification of the pupil position, which was performed using a combination of the raw SWIR images and brightness difference images. Although the eyelid skin is thicker when the eyelids are not stretched by the finger, the PLR response could be robustly identified in the new setting. Furthermore, removing the finger from the eye images reduced potential interferences due to finger movements and variability in the manner and extent of the eyelid skin stretching. Additionally, in the last experiment of the current study, the participants were not instructed to gaze forward towards a fixed point, thereby moving closer to natural bedside clinical scenarios. Despite this change, PLR responses were identified in most of the gaze directions (pupil positions) suggesting that this method can be successfully applied to monitor pupil size even when eye movements occur freely. Due to the improved imaging and analysis, the maximal PLR amplitude was greater than that observed in the first study despite the unstretched eyelids and the illumination intensity reduction due to the polarizers.

In this study we observed an apparent reversal in the intensity trends across polarization states (crossed, partially crossed, and parallel) when comparing imaging of skin (where crossed polarizers are optimal) versus the open eye (where parallel polarizers are optimal). This discrepancy may arise from differences in the optical microstructure and scattering regimes between skin and the iris. The iris may exhibit more complex birefringent and scattering behavior due to its pigment content, layered structure, and variation in scattering particle sizes. Furthermore, previous studies have reported inter-subject variability in polarization responses at visible wavelengths, suggesting that individual anatomical differences could influence the optimal polarization configuration^40,41^. Although initial data in our study hinted at such variability, the limited sample size precluded rigorous statistical analysis and warrants further investigation in future work.

Another possible explanation to the improved SNR and maximal brightness with the crossed orientation polarization when the eyelids were closed, may arise from the internal structure of the eyelids that includes vertically aligned meibomian glands embedded within the tarsal plate^51^. These elongated, lipid-rich glands may introduce anisotropic scattering that depends on both the orientation and polarization of the incident light, thus leading to the opposite trends observed between the open eyes and the closed eyes in the different polarizer configurations.

Several limitations still exist in the current study and should be acknowledged. First, although we eliminated the need to maintain a fixed gaze direction, the participants were still requested to maintain a fixed head position. At present, due to the non-uniform spatial illumination pattern of the LED, any movement could change the illumination on the eye and affect the results. Future studies should ensure a more uniform illumination to overcome this challenge. Including multiple light sources can also minimize shadows that can affect measurements. Beyond illumination, a tracking system can be implemented to allow for continuous imaging during head movements, as in the case of natural sleep. It should be noted however, that applications for anesthesia or sedation monitoring could be implemented under the assumption that head movements are minimal. Software tools can also be used to partially overcome translation and stabilize eye position in the frame. Second, due to the long duration of each trial, only a limited number of polarizer configurations could be tested. While the crossed configuration generally reduced glare, individual differences were observed. This suggests that the optimal configuration may vary between individuals, highlighting the potential value of personalization in future implementations. As the polarizers allow for better pupil position identification, it may be beneficial in some cases to use a polarizer configuration for the identification, while observing the PLR signal through a configuration that does not include polarizers. Third, the study was conducted on a relatively small number of participants. To compensate for the relatively small sample size, ten PLR events and both eyes were included in the analysis, bringing the total number of samples to 120 for each configuration.

Further research is required to demonstrate the validity of this method for identifying changes in pupil size that are smaller in magnitude than the PLR, and changes that occur in unknown times (not triggered by external stimuli). Such changes could occur due to internet state, depth of anesthesia, pain, arousal, or cognitive processes. Next steps will include experiments to monitor smaller pupillary changes, such as in response to pain or auditory stimuli, and estimate the threshold of detection at different scenarios. Additionally, future studies with large-scale data could also employ machine-learning models to predict pupil size and position from the closed eye images, along the lines of our previous study^28^. Such models can potentially better overcome noise and variability compared with the analysis approach presented here.

## Conclusion

Closed eye pupillometry can benefit from the use of polarizers to reduce glare and improve pupil position identification. The use of the crossed polarizer orientation allows better identification of the pupil position during PLR events, which enables improved PLR detection and gaze direction in naturally closed eyes.

## Data Availability

All data produced in the present work are contained in the manuscript

